# Contributing Factors to Advanced Brain Aging in Depression and Anxiety Disorders

**DOI:** 10.1101/2020.06.16.20132613

**Authors:** Laura K.M. Han, Hugo G. Schnack, Rachel M. Brouwer, Dick J. Veltman, Nic J.A. van der Wee, Marie-José van Tol, Moji Aghajani, Brenda W.J.H. Penninx

## Abstract

Brain aging has shown to be more advanced in patients with Major Depressive Disorder (MDD). This study examines which factors underlie this older brain age. Adults aged 18-57 years from the Netherlands Study of Depression and Anxiety underwent structural MRI. A pre-trained brain age prediction model based on >2,000 samples from the ENIGMA consortium was applied to predict age in 65 controls and 220 patients with current MDD and/or anxiety disorder. Brain-predicted age differences (brain-PAD) were calculated (predicted brain age minus chronological age) and associated with clinical, psychological, and biological factors. After correcting for antidepressant use, brain-PAD was significantly higher in MDD (+2.78 years) and anxiety patients (+2.91 years) compared to controls. Findings further indicate unique contributions of higher severity of somatic depression symptoms to advanced brain aging and a potential protective effect of antidepressant medication (-2.53 years).

## INTRODUCTION

Depression and anxiety are common and often comorbid mental health disorders, and their effects can broadly impact a person’s life. There is a plethora of evidence showing poorer quality of life, functional disability, and increased mortality burden in these patients (Bijl & Ravelli, 2000; Walker et al., 2015). Depression and anxiety disorders further represent a risk factor for aging-related conditions (Penninx et al., 2013; Révész et al., 2016; Verhoeven et al., 2015), as studies show consistent evidence for poorer somatic and chronic disease profiles in these patient groups (Roy-Byrne et al., 2008), often with a premature onset. Importantly, the incidence and burden of these disorders are a strain on society, which has an important challenge to face in the coming years, as the number of people aged >65 is expected to reach 1.6 billion in 2050 (Ferrucci et al., 2019). Advancing mental health and well-being across the lifespan and into old age should, therefore, be a major priority on the research agenda.

Multivariate pattern recognition techniques, and especially machine learning methods, have promoted a steep increase in the development of ways to measure and quantify aging (Jylhava et al., 2017). Central to this field is that multivariate (biological) patterns are utilized and integrated into a single score: the biological age. Biological age can be derived from, for instance, omics-data (e.g. epigenetic clocks), but also clinical biomarkers obtained from, for example, blood chemistries (Hägg et al., 2019). In the current study, we focus on biological age based on a validated method of MRI-derived brain structure (Cole & Franke, 2017; Gaser & Franke, 2019) with brain-predicted age difference (brain-PAD, predicted brain age minus chronological age) (Cole et al., 2019) as the main outcome. This metric is relative to one’s chronological age, such that positive values indicate an older appearing brain, and negative values resemble a younger appearing brain than normally expected at that age.

A handful of studies have investigated brain-PAD in depression, with studies showing +4.0 years (Koutsouleris et al., 2014), as well as no significantly increased brain age (Besteher et al., 2019; Kaufmann et al., 2019). Recent findings from the Enhancing NeuroImaging Genetics through Meta-Analysis (ENIGMA) consortium using a more than ten-fold larger pooled sample of MDD patients than the largest previous study suggest a 1.1 year higher brain-PAD in MDD patients as compared to controls (Han et al., 2019).

However, this difference did not seem to be driven by specific clinical characteristics (recurrent status, remission status, antidepressant medication use, age of onset, or symptom severity). An important aspect that remains relatively unknown is thus which underlying mechanisms cause the brain age metric to advance in depression, and, despite the increase of brain age studies in the past decade, in general (Gaser & Franke, 2019). Large pooled datasets from global consortia offer the statistical power needed to detect small effect sizes usually observed in MDD, but a limitation of consortium data is that its collection is commonly not harmonized across all sites and cohorts. Here, we underline the complementary value of a more homogeneous and clinically well-characterized sample from the Netherlands Study of Depression and Anxiety (NESDA), to gain more insight into the observed brain-PAD difference between MDD patients and controls. We extend prior work by exploring which specific symptom clusters (mood/cognition, immunometabolic, somatic) of MDD are associated with brain-PAD. To the best of our knowledge, there are currently no brain age studies in anxiety disorders, although higher brain-PAD has been observed in posttraumatic stress disorder (Liang et al., 2019). Given the frequent co-occurrence and correlated symptoms (Krueger & Markon, 2006) of depression and anxiety (i.e. family of internalizing disorders) (Caspi et al., 2020), we also extend prior work by including patients with MDD and/or anxiety disorders in the current study.

Evidence is starting to emerge that brain-PAD is associated with reduced mental and somatic health, such as with stroke history, diabetes diagnosis, smoking, alcohol consumption, and some cognitive measures (Cole, 2020), but also intrinsic measures such as genetic variants (Jonsson et al., 2019; Ning et al., 2020). This study seeks to further address the research gaps, by examining whether three important biological stress systems that are commonly found to be dysregulated in depression and anxiety disorders (inflammation, hypothalamic pituitary adrenal [HPA]-axis, autonomic nervous system [ANS]), were predictive of brain aging. Disruptions and dysregulations in these stress systems were hypothesized to result in advanced brain aging across diagnostic groups. We further associated various clinical, lifestyle, and somatic health indicators with the brain-PAD metric to identify unique contributing factors to brain aging.

## RESULTS

### Sample Characteristics

Demographics and assessed phenotypes of the current study sample can be found in **Table 1**. Briefly, the patient group consisted of patients with a current MDD diagnosis but no anxiety (28.2%), patients with a current anxiety disorder but no depression (30.5%), and patients with a current comorbid depression and anxiety disorder (41.4%). The patient group (mean 37.37 ± SD 10.20 years) was younger than the control group (mean 40.81 ± SD 9.78 years) and had fewer years of education (mean 14.28 ± SD 2.86 years in controls vs. mean 12.39 ± SD 3.19 in patients). Control and patient groups were similar in terms of male/female ratios, but not distributed equally between scan locations (Amsterdam, Leiden, Groningen) (***X***_(2)_=6.26, p=0.044).

**Table 1:** Participant Characteristics of Controls and Patients.

| Characteristic | N | Controls, N = 65<sub>1</sub> | Patients, N = 220<sub>1</sub> | p-value<sub>2</sub> |
| --- | --- | --- | --- | --- |
| *Demographics* |  |  |  |  |
| Age (years) | 285 | 40.81 ± 9.78 (21.26-56.67) | 37.37 ± 10.20 (17.76-57.17) | **0.02** |
| Female Sex | 285 | 42 (65%) | 152 (69%) | 0.60 |
| Education Level (years) | 285 | 14.28 ± 2.86 (5.00-18.00) | 12.39 ± 3.19 (5.00-18.00) | **<0.001** |
| Scanlocation | 285 |  |  | **0.04** |
| 1 |  | 26 (40%) | 66 (30%) |  |
| 2 |  | 27 (42%) | 78 (35%) |  |
| 3 |  | 12 (18%) | 76 (35%) |  |
| *Clinical characteristics* |  |  |  |  |
| Major depressive disorder |  |  | 62 (28%) |  |
| Anxiety disorder |  |  | 67 (30%) |  |
| Comorbid depression and anxiety |  |  | 91 (41%) |  |
| Total depression severity score | 280 | 4 ± 4 (0-21) | 23 ± 12 (1-57) | **<0.001** |
| Total anxiety severity score | 278 | 2 ± 3 (0-11) | 14 ± 10 (0-50) | **<0.001** |
| Mood/cognition symptom cluster | 285 | 1.09 ± 0.13 (1.00-1.47) | 1.86 ± 0.50 (1.00-3.27) | **<0.001** |
| Somatic depression symptom cluster | 285 | 1.20 ± 0.21 (0.90-2.20) | 1.64 ± 0.41 (0.80-2.80) | **<0.001** |
| Immunometabolic symptom cluster | 285 | 1.11 ± 0.21 (0.80-1.80) | 1.60 ± 0.48 (0.60-3.60) | **<0.001** |
| Childhood Trauma Index | 285 | 1 ± 1 (0-8) | 2 ± 2 (0-8) | **<0.001** |
| Recent negative life events | 285 | 0.57 ± 0.83 (0.00-3.00) | 0.89 ± 1.09 (0.00-3.00) | 0.05 |
| *Within patients* |  |  |  |  |
| Antidepressant use | 220 |  | 77 (35%) |  |
| Duration of depressive symptoms<br>(proportion of time in the past 4 years) | 190 |  | 0.34 ± 0.28 (0.00-1.00) |  |
| Duration of anxiety symptoms<br>(proportion of time in the past 4 years) | 192 |  | 0.42 ± 0.35 (0.00-1.00) |  |
| Age of onset of depression (years) | 191 |  | 23.75 ± 10.44 (4.00-54.00) |  |
| Age of onset of anxiety (years) | 170 |  | 18.15 ± 10.93 (4.00-52.00) |  |
| *Somatic health* |  |  |  |  |
| Body Mass Index (kg/m<sup>2</sup>) | 285 | 24.36 ± 3.73 (19.03-37.42) | 25.14 ± 4.72 (18.04-42.21) | 0.35 |
| Number of somatic diseases | 285 | 0 ± 1 (0-3) | 0 ± 1 (0-3) | 0.66 |
| *Lifestyle* |  |  |  |  |
| Alcohol intake (mean number of drinks per week) | 285 | 6.2 ± 6.1 (0.0-25.0) | 4.3 ± 6.5 (0.0-47.5) | **0.01** |
| Smoking behavior (cigarettes/day) | 161 | 9.26 ± 7.61 (0.00-29.00) | 12.56 ± 10.73 (0.00-70.00) | 0.09 |
| Physical activity (1,000 MET minutes per week) | 271 | 3.8 ± 3.3 (0.3-16.5) | 3.6 ± 3.5 (0.0-17.1) | 0.19 |
| *Inflammation* |  |  |  |  |
| C-Reactive Protein (mg/l) | 280 | -0.03 ± 0.52 (-1.00-1.08) | 0.11 ± 0.59 (-1.00-1.35) | 0.17 |
| Tumor Necrosis Factor-α (pg/ml) | 279 | -0.16 ± 0.26 (-1.00-0.63) | -0.12 ± 0.26 (-1.00-0.63) | 0.39 |
| Interleukin-6 (pg/ml) | 280 | -0.15 ± 0.31 (-1.12-0.57) | -0.14 ± 0.48 (-2.29-1.74) | 0.89 |
| *Autonomic Nervous System* |  |  |  |  |
| Resting Heart Rate (bpm) | 276 | 69 ± 8 (51-86) | 68 ± 10 (44-96) | 0.62 |
| Respiratory Sinus Arrhythmia (ms) | 276 | 51 ± 25 (14-130) | 49 ± 26 (7-130) | 0.59 |
| Pre-injection Period (ms) | 276 | 119 ± 17 (81-147) | 119 ± 16 (75-168) | 0.66 |
| *HPA-axis* |  |  |  |  |
| Cortisol Awakening Response Area under the curve with respect to the ground (nmol/l/hr) | 197 | 0.57 ± 4.63 (-13.06-11.13) | 2.44 ± 5.74 (-14.92-19.00) | 0.10 |
| Cortisol Awakening Response Area<br>under the curve with respect to the<br>increase (nmol/l/hr) | 197 | 16.89 ± 4.88 (8.49-32.13) | 18.51 ± 6.75 (5.36-37.97) | 0.23 |
| Evening cortisol (nmol/l) | 207 | 5.05 ± 2.44 (2.12-13.33) | 5.04 ± 2.43 (1.09-12.96) | 0.90 |

<sup>1</sup> Statistics presented: mean ± SD (minimum-maximum); n (%)
<sup>2</sup> Statistical tests performed: Wilcoxon rank-sum test; chi-square test of independence

### Brain age prediction performance

Using the ENIGMA brain age model (www.photon-ai.com/enigma_brainage) we obtained a correlation of r=0.73 in the control subjects and r=0.72 in the patient group between predicted and chronological age, but in both groups brain age predictions were overestimated (mean brain-PAD [SD]; 8.18 [7.27] years in controls and 10.86 [7.73] years in patients). To correct for the offset, we calculated the mean brain-PAD in the control group and subtracted these from all individual brain-PAD estimates. This correction resulted in an R_2_ of 0.45 and MAE of 5.97 (SD 4.09) years in controls, and R_2_ of 0.36 and MAE of 6.73 (4.64) years in patients (also see **Table 2** for model metrics). Of note, this linear correction does not affect subsequent statistics. **Figure 1A** shows the unaffected correlation between predicted brain age (x-axis) and chronological age (y-axis) in control subjects (r=0.73, p<0.0001) and in patients (r=0.72, p<0.0001). There also was a well-known and commonly described age-bias (i.e. correlation between brain-PAD and age) (Le, Kuplicki, McKinney, et al., 2018; Liang et al., 2019; Smith et al., 2019) in controls (r=-0.32, p=0.01) and patients (r=-0.37, p<0.0001) in the current sample (**Figure 1B)**, which was statistically dealt with by including age as a predictor variable in further analyses (**Figure 1C)** (Le, Kuplicki, McKinney, et al., 2018).

**Table 2:** Model metrics.

|  | r | R2 | MAE | SD | RMSE | Brain-PAD | SD |
| --- | --- | --- | --- | --- | --- | --- | --- |
| Controls | 0.73 | -0.26 | 9.01 | 6.19 | 10.91 | 8.18 | 7.27 |
| Controls corrected | 0.73 | 0.45 | 5.97 | 4.09 | 7.22 | 0 | 7.27 |
| Patients | 0.72 | -0.71 | 11.38 | 6.95 | 13.32 | 10.86 | 7.73 |
| Patients corrected | 0.72 | 0.36 | 6.73 | 4.64 | 8.17 | 2.69 | 7.73 |

Performance metrics of the ENIGMA model applied to the NESDA data per diagnostic group. r, Pearson's correlation coefficient; R<sub>2</sub>, proportion of explained variance; MAE, mean absolute error in years; SD, standard deviation; RMSE, root mean squared error; Brain-PAD, mean brain predicted age difference in years (predicted age - actual age). Corrected metrics reflect brain-PAD estimates - mean brain-PAD<sub>controls</sub> .

**Figure 1:**
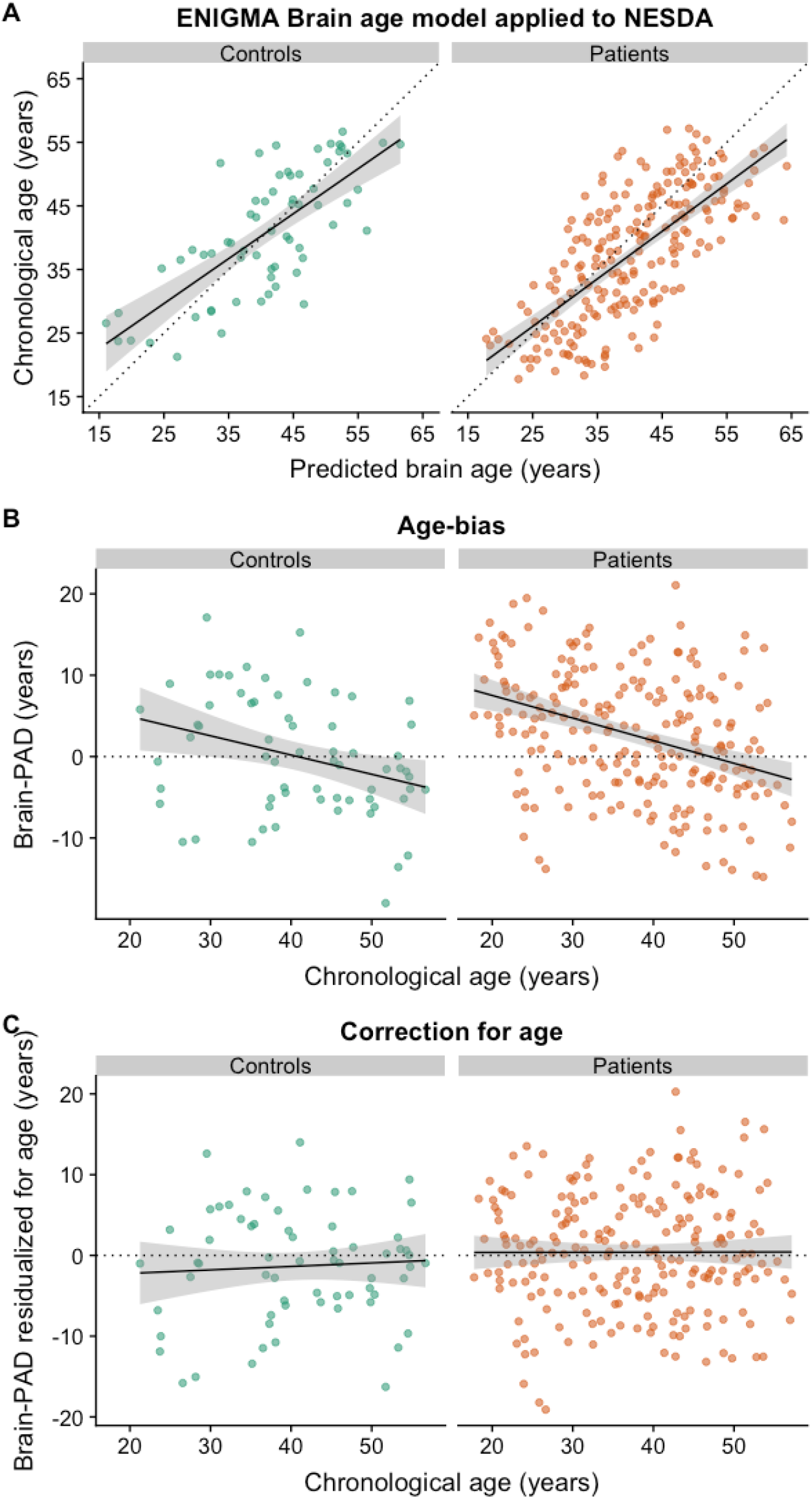
Brain age prediction. **(A)** Correlation between predicted brain age and chronological age in controls (r=0.73, R_2_=0.45, p<0.0001) and patients (r=0.72, R_2_=0.36, p<0.0001). Of note, predicted brain age reflects estimates corrected for the offset (brain age_corrected_ = brain age - (brain-PAD - mean brain-PAD_controls_). **(B)** There was a residual effect of age on the brain-PAD outcome in controls (r=-0.32, p=0.01) and patients (r=-0.37, p<0.0001), **(C)** which was statistically corrected for by adding age as a covariate in all models.

### Advanced brain aging in depression and anxiety disorders

Using diagnostic status as a dichotomous between-group predictor we found that patients exhibited +1.75 years higher brain-PAD than controls, but this difference did not reach statistical significance (Cohen’s d=0.24). Within the patient group only, we found no significant associations with the age of onset of illness or duration of symptoms of either MDD or anxiety. However, brain-PAD was significantly lower in antidepressants (AD) using patients compared to AD-free patients (b=-2.58 years, p=0.01), but not control subjects (b=0.59 years, p=0.65) (**Figure 2**). Given the significant difference in brain-PAD between AD-free and AD-using patients, we included AD status as an additional covariate when comparing controls to the patient group, resulting in significantly higher brain-PAD in patients (+2.63 year [SE 1.10 year], Cohen’s d=0.34, 95% CI 0.06-0.62). We also added AD status as an additional covariate in a model to compare controls against specific MDD, anxiety, or comorbid patient groups (the proportion of subjects using AD in specific diagnostic groups was marginally different, ***X***_(2)_=5.91, p=0.052). This revealed significantly higher brain-PAD in MDD (+2.78 years, Cohen’s d=0.25, 95 % CI -0.10-0.60, p=0.04) and anxiety patients (+2.91 years, Cohen’s d=0.27, 95% CI -0.08-0.61, p=0.03), and a similar effect in the comorbid MDD and anxiety group (+2.23 years, Cohen’s d=0.21, 95% CI 0.10-0.53) although only marginally significant (p=0.08) (**Table 3**). There were no post-hoc differences in brain-PAD corrected for AD use between specific patient groups (MDD vs. anxiety vs. comorbid patients; P’s>0.46, Cohen’s d’s <0.07).

**Table 3:** Advanced brain aging in depression and anxiety with correction for antidepressant use.

| Ref | Predictor | b | SE | t value | P | Cohen's d | SE | 95% CI |
| --- | --- | --- | --- | --- | --- | --- | --- | --- |
| Controls | Any patient | 2.63 | 1.10 | 2.39 | 0.02 | 0.34 | 0.14 | 0.06-0.62 |
|  | MDD | 2.78 | 1.32 | 2.11 | 0.04 | 0.25 | 0.18 | -0.09-0.6 |
|  | Anxiety | 2.91 | 1.31 | 2.22 | 0.03 | 0.27 | 0.17 | -0.08-0.61 |
|  | Comorbid MDD and anxiety | 2.23 | 1.28 | 1.74 | 0.08 | 0.21 | 0.16 | -0.11-0.53 |

Age, sex, education level (years) and two dummy variables for scanlocation were included in all models. Antidepressant status was additionally included as covariate.

**Figure 2:**
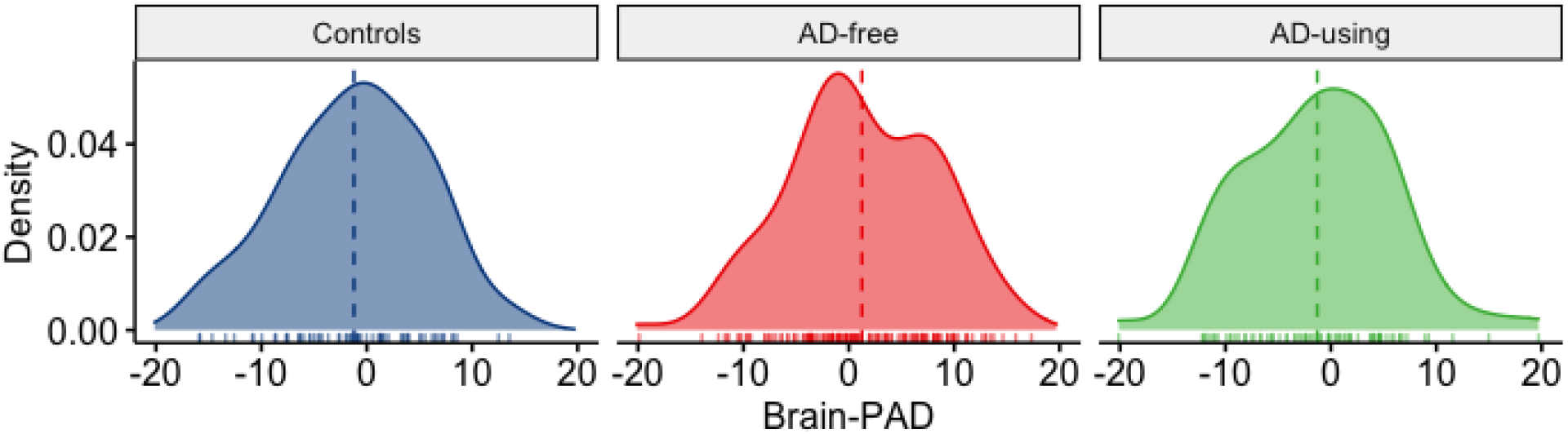
Brain-PAD differences between controls, antidepressant using (AD-using) and antidepressant-naive (AD-free) patients. AD-free patients showed significantly higher brain-PAD compared to AD-using patients (+2.58 year [SE 1.02 year], Cohen’s d=0.36, 95% CI 0.09-0.64) and controls (+2.63 year [SE 1.10 year], Cohen’s d=0.31, 95% CI 0.01-0.60). Brain-PAD estimates (in years) were residualized for age, sex, education level (years) and two dummy variables for scanlocation.

To gain more insight into the differences in brain-PAD between AD-free and AD-using patients, we post-hoc calculated a derived daily dose by dividing the AD mean daily dose by the daily dose recommended by the World Health Organization (also see (Licht et al., 2008)). Brain-PAD was not significantly negatively associated with a derived daily dose of antidepressants in n=74 patients (b=-0.91 year, p=0.50) (**Supplementary Figure S1**). Of note, we excluded three subjects from this analysis as these AD-using patients were using Venlafaxine at doses higher than 150 mg/day, acting as a dual serotonin and norepinephrine reuptake inhibitor rather than acting as a Selective Serotonin Reuptake Inhibitor (SSRI) only (Debonnel et al., 2007). Based on the above findings, both diagnostic and AD status were included in the multivariate model to test unique brain-PAD contributions.

### Selection of significant associations with clinical variables in all participants

Using a dimensional approach based on symptoms rather than diagnosis, we found that higher brain-PAD was associated with higher total depression (b=0.07 year per unit change on the Inventory of Depressive Symptoms, p=0.03) and anxiety severity scores (b=0.11 year per unit change on the Beck’s Anxiety Inventory, p=0.01) across all participants (**Figure 3A-B**). No significant associations were found for the mood/cognition (b=0.89 year per unit increase on the average sum score, p=0.27, **Figure 3C**) or immunometabolic symptom clusters of depression (b=0.45 year per unit increase on the average sum score, p=0.62, **Figure 3D**), but higher brain-PAD was strongly associated with more somatic symptoms of depression (b=4.03 year per unit increase on the average sum score, p<0.0001, **Figure 3E**). There were no significant associations between brain-PAD and childhood trauma exposure (b=0.23 year per unit change on the childhood trauma index, p=0.26) or recent negative life events (b=0.35 year per negative life event, p=0.39).

**Figure 3:**
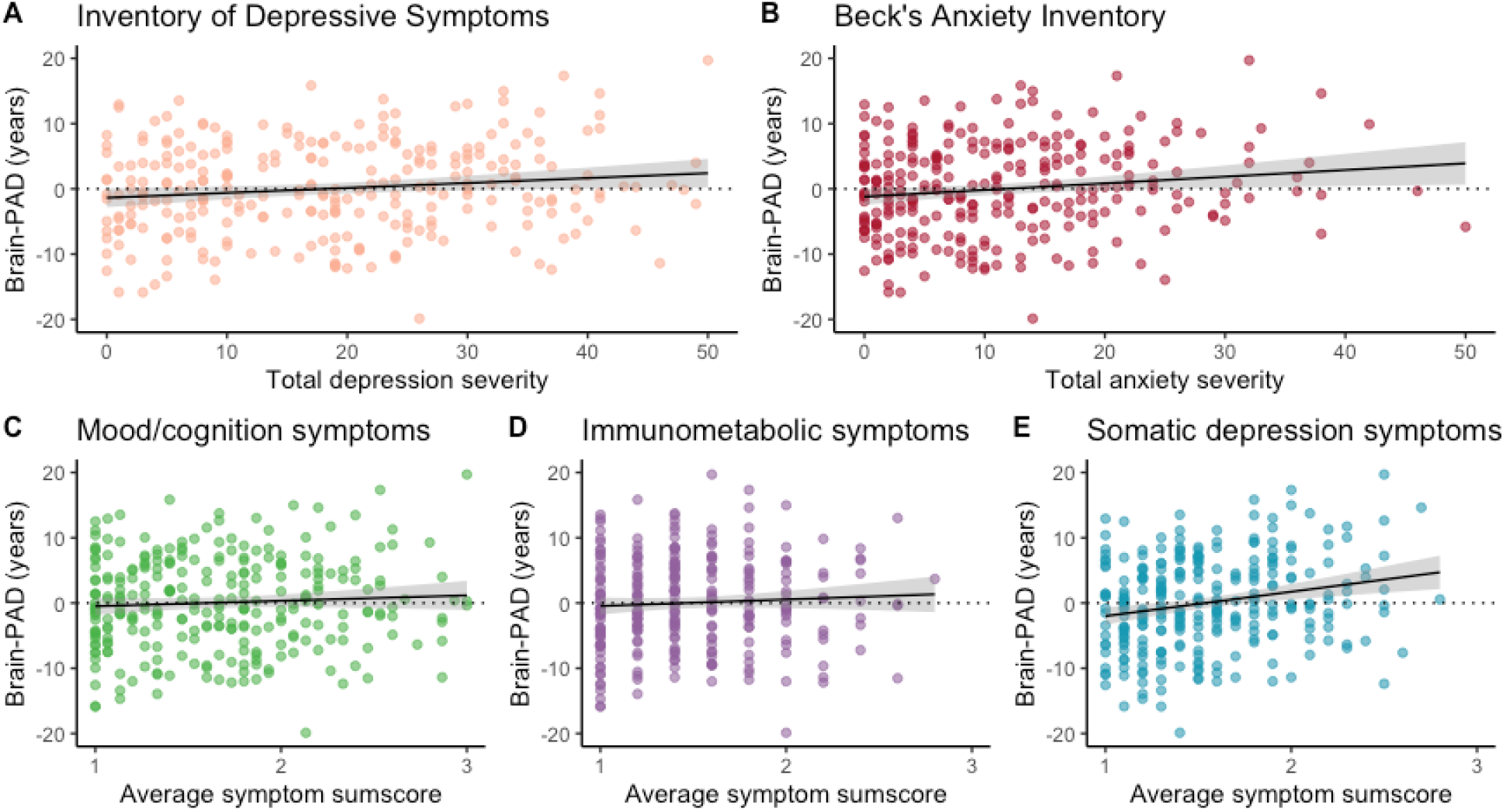
Brain-PAD associations with symptom severity. **(A)** Advanced brain aging was associated with overall higher total depressive symptoms (b=0.07 years per unit increase on the Inventory of Depressive Symptoms, p=0.03), **(B)** total anxiety symptoms (b=0.11 years per unit increase on the Beck’s Anxiety Inventory, p=0.01), but not specifically with **(C)** the mood/cognition (b=0.89 years per unit increase on average sum score, p=0.27) or **(D)** immunometabolic (b=0.45 years per unit increase on the average sum score, p=0.62) symptom cluster. The association in (A) seemed to be driven mostly by **(E)** a specific cluster of somatic symptoms in MDD (b=4.03 years per unit increase on the average sum score, p<0.0001). Brain-PAD estimates (in years) were residualized for age, sex, education level (years) and two dummy variables for scanlocation.

### Selection of significant associations with somatic health in all participants

Higher brain-PAD was associated with both higher BMI (b=0.23 year per kg/m_2_, p=0.02, **Figure 4**), as well as the number of somatic diseases under medical treatment (b=1.45 year per somatic disease, p=0.03). However, the latter association became non significant if those with >2 chronic diseases (n=4) were truncated to two chronic diseases (b=1.29 year per somatic disease, p=0.08).

**Figure 4:**
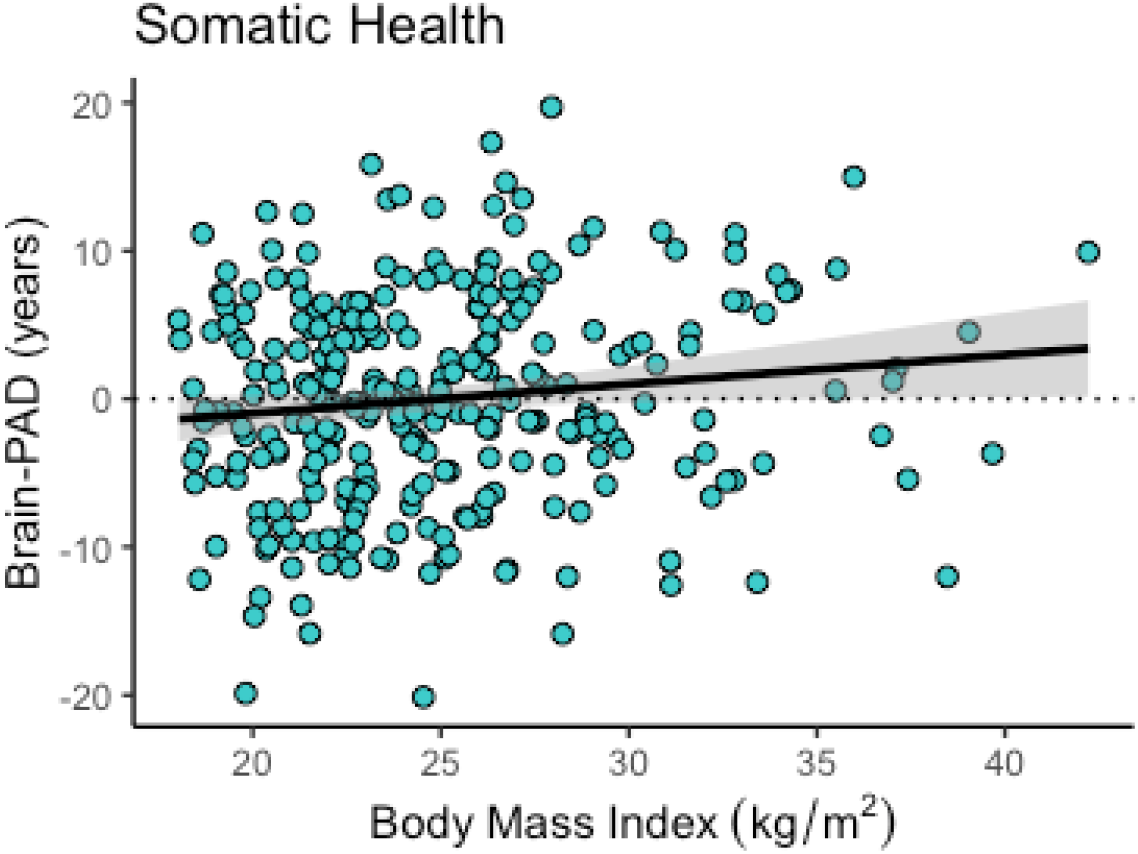
Brain-PAD associations with somatic health. Each increase of 1 kg/m_2_ in BMI leads to +0.23 years of added brain aging (p=0.02). Brain-PAD estimates (in years) were residualized for age, sex, education level (years) and two dummy variables for scanlocation.

### No associations with lifestyle or biological stress variables

There were no significant associations with any of the lifestyle variables (smoking, alcohol, physical activity) or biological stress variables (inflammatory markers, ANS, HPA-axis). An overview of the separate linear regressions can be found in **Table 4**.

**Table 4:** Overview of the brain-PAD associations with predictors of interest.

| **Assessment** | **Predictor** | **b** | **SE** | **t value** | **P** |
| --- | --- | --- | --- | --- | --- |
| *Clinical* | Depressive symptom severity | 0.07 | 0.03 | 2.16 | **0.03** |
|  | Anxiety symptom severity | 0.11 | 0.04 | 2.50 | **0.01** |
|  | Mood/cognition symptoms | 0.89 | 0.81 | 1.10 | 0.27 |
|  | Somatic depression symptoms | 4.03 | 1.04 | 3.87 | **<0.00001** |
|  | Immunometabolic symptoms | 0.45 | 0.92 | 0.49 | 0.62 |
|  | Childhood trauma index | 0.23 | 0.20 | 1.13 | 0.26 |
|  | Negative life events | 0.35 | 0.41 | 0.86 | 0.39 |
| *Within patients* | Antidepressant use | -2.58 | 1.02 | -2.54 | **0.01** |
|  | Duration of depressive symptoms | -0.20 | 1.97 | -0.10 | 0.92 |
|  | Duration of anxiety symptoms | -0.88 | 1.55 | -0.56 | 0.57 |
|  | Age of onset of depression | 0.04 | 0.06 | 0.61 | 0.55 |
|  | Age of onset of anxiety | 0.01 | 0.05 | 0.09 | 0.93 |
| *Somatic health* | BMI | 0.23 | 0.10 | 2.31 | **0.02** |
|  | Number of somatic diseases | 1.29 | 0.72 | 1.79 | 0.08 |
| *Lifestyle* | Alcohol (mean drinks per week) | -0.09 | 0.07 | -1.33 | 0.19 |
|  | Smoking (cigarettes per day) | -0.07 | 0.05 | -1.26 | 0.21 |
|  | Physical exercise (MET-minutes) | -0.06 | 0.13 | -0.48 | 0.63 |
| *Inflammation* | CRP | 0.57 | 0.76 | 0.75 | 0.46 |
| | TNF-α | 0.10 | 1.65 | 0.06 | 0.95 |
|  | IL6 | 0.60 | 0.98 | 0.61 | 0.54 |
| ANS | Resting HR | 0.08 | 0.05 | 1.51 | 0.13 |
|  | RSA | -0.01 | 0.02 | -0.46 | 0.65 |
|  | PEP | -0.04 | 0.03 | -1.45 | 0.15 |
| HPA-axis | AUCi | -0.04 | 0.10 | -0.34 | 0.74 |
|  | AUCg | -0.04 | 0.09 | -0.42 | 0.67 |
|  | Evening | 0.18 | 0.23 | 0.76 | 0.45 |

Age, sex, education level (years) and two dummy variables for scanlocation were included in all models. BMI, Body Mass Index; MET-minutes, ;CRP, C-reactive protein; TNF-α , Tumor Necrosis Factor- $\alpha$ ; IL6, Interleukin-6; ANS, autonomic nervous system; HR, heart rate; RSA, respiratory sinus arrhythmia; PEP, pre-ejection period; AUCi, cortisol awakening response: area under the curve with respect to the increase; AUCg, cortisol awakening response: area under the curve with respect to the ground; Evening, Evening Cortisol.

### Multivariable model

To characterize the unique contributions of the selected significant predictors on the brain-PAD outcome, we included diagnostic status (control vs. patient), MDD and anxiety symptom scores, BMI, AD use, and the number of somatic diseases under treatment as predictors in a stepwise regression model with forward selection. Thus, predictors were successively added to an intercept-only model (Akaike’s Information Criterion [AIC] = 1115.81), only adding regression coefficients if they improved model fit (i.e. lower AIC). Using this method, we found that the best subset of variables to explain brain-PAD consisted of somatic depression symptoms and AD use (AIC=1098.79). In sum, unique contributions to brain-PAD were observed for the somatic depression symptom cluster (b=4.21 year per unit increase on average sum score, p<0.0001) and AD use (b=-2.53 year, p=0.007) **(Table 5**).

**Table 5:** Result of stepwise regression model with forward selection of brain-PAD contributors.

| N | Selected predictors | b | 95% CI | SE | t value | P |
| --- | --- | --- | --- | --- | --- | --- |
| 285 | Somatic depression symptoms | 4.21 | 2.25 to 6.16 | 1.00 | 4.22 | **<0.0001** |
|  | AD Use | -2.53 | -4.36 to -0.70 | 0.93 | -2.72 | **0.007** |

Current model with lowest AIC=1098.79 compared to intercept-only model AIC=1115.81. The brain-PAD outcome was residualized for age, sex, educational level (years) and two dummy variables for scanlocation.

## DISCUSSION

The current study used a validated brain age prediction model to show that the previously observed findings of older appearing brains in MDD patients was associated with symptom severity and BMI. Moreover, antidepressant (AD) users exhibited similar average brain-PAD as control subjects, whereas those that were AD-free showed older appearing brains. Correcting for AD-use, we also showed that not only MDD patients, but also patients with anxiety disorder exhibited older appearing brains compared to controls. Surprisingly, there were no significant associations with lifestyle or biological stress systems. A multivariable model showed unique contributions of somatic depression symptom severity and AD-use on brain-PAD.

To the best of our knowledge, we are the first to report advanced brain aging in anxiety disorders (i.e. generalized anxiety disorder, panic disorder, social anxiety disorder) with an estimated +2.91 years on average, compared to controls, when correcting for AD use. This is consistent with the literature describing comparable effect sizes with respect to structural brain alterations in social anxiety disorder (Cohen’s d=0.20) (Bas-Hoogendam et al., 2017), and other anxiety-related disorders such as post-traumatic stress disorder (PTSD) (Cohen’s d=-0.17) (Logue et al., 2018), with PTSD patients also showing advanced brain-PAD without correction for AD (Liang et al., 2019). This observation may potentially offer an explanation as to why clinical anxiety is associated with an increased risk of dementia, even independent from depression (Santabárbara et al., 2019), although further evidence is needed. The lack of any significant post-hoc differences between specific diagnostic groups can likely be explained due to, amongst others, the high genetic correlation between the disorders (Brainstorm Consortium et al., 2018), shared environmental risks, and overlapping personality traits of patients with depression and anxiety disorders (Ruscio & Khazanov, 2017).

The most clinically relevant finding was that AD-using patients showed a similar brain age to controls, but not to AD-free patients, irrespective of specific depressive or anxiety disorder. This finding was previously overlooked in consortium data, presumably due to a lack of more detailed information on lifetime use, dosage and duration of use of AD (Han et al., 2019), highlighting the complementary values of well-characterized local samples and large-scale consortia. The AD finding was particularly interesting as the AD-using patients constituted a more severely depressed and anxious group as indicated by higher symptom severities compared to AD-free patients, potentially suggesting compensatory or normalizing mechanisms of AD, at least on the brain-PAD metric. This accords with earlier work reporting brain-PAD associations with therapeutic drugs, suggesting neuroprotective effects of Lithium treatment in bipolar disorder patients (vs. no Lithium) (Van Gestel et al., 2019) and ibuprofen (vs. placebo) in healthy participants in a exploratory randomized controlled trial (Le, Kuplicki, Yeh, et al., 2018). Yet, it remains unclear if and to what extent the brain age protective mechanisms overlap with, for example, increased neural progenitor cells (Boldrini et al., 2012), brain-derived neurotrophic factor (BDNF) (Castrén & Kojima, 2017), or other serotonergic neuroplasticity processes implicated in AD use (Kraus et al., 2017), or, alternatively, whether neuropharmacology affects the MRI signal (Cousins et al., 2013). Brain-PAD was not positively associated with the duration of symptoms (either MDD or anxiety), suggesting that the AD effect was not driven by the duration of the disease and did not seem to be progressive. Taken together, these findings may suggest an age-related neuroprotective effect of AD, but interpretative caution is warranted as the current study was cross-sectional in nature and the dose-response association with AD not statistically significant. We also did not find associations with physical activity, while a previous study found an association between brain-PAD and the daily number of flights of stairs climbed (Steffener et al., 2016). Future clinical interventions are needed to examine the short and long-term effects of antidepressants and physical activity on biological aging, an objective currently pursued by the MOod treatment with antidepressants or running (MOTAR) study (Lever-van Milligen et al., 2019).

There were no associations with the cumulative childhood trauma index or the number of recent negative life events, different from the impact that adverse childhood experience commonly has on other biological age indicators such as telomere length (Aas et al., 2019), or epigenetic aging (Wolf et al., 2018), albeit with small effects. Future studies with larger samples may potentially be more sensitive in picking up associations between brain-PAD and childhood trauma. However, taken together, the current study found that advanced brain aging was more associated with current disease states, likely related to current symptom severity, rather than the result of cumulative exposure (i.e. no association with childhood trauma history, age of onset of illness, duration of symptoms) or traits.

Furthermore, Cole and colleagues (2020) found significant associations between brain-PAD and several biomedical (e.g. blood pressure, diabetes, stroke) and lifestyle variables (e.g. smoking status, alcohol intake frequency), but not BMI, in the UK Biobank (Cole, 2020), albeit with a different, multi-modal brain age prediction model but in a much larger sample size (>14,000 subjects). Although the current findings with somatic health broadly support previously associated diabetes (Franke et al., 2013) and stroke findings in UK Biobank, as well as the null-finding with respect to physical activity, we did not identify associations with smoking or alcohol behavior (Franke et al., 2013). More work is needed in terms of identifying unique or shared robust contributors to the brain-PAD metric, converging evidence across and between datasets, processing methods, and populations. Other previous studies, however, also identified associations with BMI (Franke et al., 2014) and here we show that an increase of 1 kg/m_2_ in BMI leads to +0.23 years of added brain aging, although not independent from depression or anxiety symptom severity. A previous study did show such an independent effect for obesity and first-episode schizophrenia (Kolenic et al., 2018), but here the obese group only constituted of 13% of the total current sample. Furthermore, each increase of 1 of the average sum score (range 0.80-4.00) of somatic depression symptoms, resulted in +4.20 years of added brain aging, independent from AD use. The somatic symptom cluster studied here consisted of items tapping into sleep, psychomotor, and other bodily symptom problems (see **Supplement** for all individual items within each cluster). This emphasizes the need to prevent and improve both mental and somatic conditions to promote healthy brain aging in psychiatric populations.

Surprisingly, none of the biological stress systems considered in the current study were predictive of brain aging, despite the strong association between brain-PAD and somatic symptoms. This suggests that the biological dysregulations that commonly link depression to somatic health (Penninx et al., 2013), were not directly contributing to advanced brain aging. On the other hand, it might indicate that the brain-PAD metric is more responsive to psychological stressors, rather than biological stressors. With respect to the inflammatory markers, it might be possible that blood levels of inflammatory markers do not accurately mirror central neuroimmune levels, although there is some evidence that C-Reactive Protein (CRP) measured peripherally also reflects central inflammation, at least in MDD (Felger et al., 2018). Alternatively, a different potential biological mechanism that may explain the observed advanced brain aging in depression and anxiety disorders is metabolic dysregulation. Future studies could characterize the brain-PAD metric in more detail with respect to metabolic factors (e.g. blood pressure, triglycerides, cholesterol), as these are well-established risk factors for unfavorable somatic conditions (Eckel et al., 2005; Esposito et al., 2012; Mottillo et al., 2010; Profenno et al., 2010) and frequently co-occur with depression (Pan et al., 2012).

Given the richness of the current dataset, we additionally computed post-hoc intercorrelations between the brain-PAD metric and other available biological age indicators in NESDA. Briefly, we found low, non-significant (P’s>0.13), correlations between brain-PAD, and three omics-based clocks (epigenetic, transcriptomic, metabolomic) and telomere length (with Pearson *r* in the range of -0.03 to 0.15, **Supplementary Figure S2**). Surprisingly, brain-PAD was negatively associated with the proteomic clock (r=-0.24, p=0.02) after correcting for age (albeit in a greatly reduced overlapping sample of N=98). Only a handful of studies have compared multiple biological age indicators side-by-side (Belsky et al., 2018; Jansen et al., 2020; Kim et al., 2017; Murabito et al., 2018), but the current findings support most work showing the very little overlap between biological clocks from different types of data (Hägg et al., 2019). However, the small but significant negative correlation between brain and proteomic aging suggests a further study with more focus on the interplay between this peripheral and central proxy of aging is needed. Aging remains a multifaceted and complex process that may manifest differently across multiple biological levels and tissues.

### Limitations

It is important to mention that our sample had low statistical power to detect (some of) the relatively small effect sizes in the current study. At present, the large within-group variance of brain-PAD lacks utilitarian validity in a clinical context. We, therefore, emphasize the need for both methodological (i.e. brain age models) as well as epidemiological replication (i.e. other and larger samples) to test the robustness of effects. Another limitation is reflected by the lack of insights into the causal pathways implicated in advanced brain aging, given the cross-sectional nature of the study. However, a major strength is that we used a pre-established reference curve for healthy brain aging that has further potential for benchmarking, as the ENIGMA MDD working group encourages local research samples like ours to examine more detailed phenotypes that were not available within the consortium. Also important to note is that the effects of multivariate brain aging patterns (Cohen’s d=0.34, between controls and all patients) was higher or comparable to other biological aging indicators (e.g. telomere length [Cohen’s d=0.12] (Verhoeven et al., 2013), epigenetic aging [d=0.14]) (Han et al., 2018), biological markers (e.g. BDNF [d=0.23](Molendijk et al., 2011), cortisol [d=0.15-0.25] (Vreeburg et al., 2009), CRP [d=0.15] (Howren et al., 2009)), and, most importantly, neuroimaging markers (e.g. hippocampal volume [d=-0.14] (Schmaal et al., 2015)), in other or (partly) overlapping samples.

## Conclusion

In summary, advanced brain aging in patients with MDD and anxiety seems to be most strongly associated with somatic health indicators such as somatic depressive symptomatology, BMI, and the number of chronic diseases under medical treatment. We also revealed that antidepressant medication use was associated with lower brain-PAD, potentially suggesting that its use may have a protective effect on the age-related structural gray matter alterations observed in patients with MDD and anxiety, an effect previously overlooked in consortium data. Our results, therefore, emphasize the importance and complementary value of smaller, yet more homogeneous, datasets with harmonized data collection and well-characterized clinical phenotyping, compared to the large-scale consortium data needed for statistical power. Randomized clinical trials are needed to confirm whether advanced brain aging can be halted or reversed, by intervening on the cross-sectional somatic health indicators identified here, in pursuit of the characterization of a complex multifaceted process such as brain aging.

## METHODS AND MATERIALS

### Subjects

A subsample of subjects of the Netherlands Study of Depression and Anxiety (NESDA) were included for the MRI substudy (total N=301). Twelve participants were excluded due to poor image quality, two because of claustrophobia, one control subject due to high depression rating (Montgomery Asberg Depression Rating Scale score >8), and one due to the large time difference between the psychiatric and biological and MRI measurements (total excluded, N=16). For the current study, we therefore included N=65 controls (65% female, aged 21-55) and N=220 patients with a current depressive and/or anxiety disorder (69% female, aged 18-57). The current study was approved by the ethical review boards of the three participating centers (Amsterdam, Groningen, Leiden) and informed consent of all participants was obtained.

### Image processing and analysis

Magnetic resonance imaging (MRI) data were obtained using three independent 3T Philips MRI scanners (Philips Healthcare, Best, The Netherlands) located at different participating centers. Scanners were equipped with a SENSE 8-channel (Leiden University Medical Center and University Medical Center Groningen) and a SENSE 6-channel (Academic Medical Center) receiver head coil (Philips Healthcare). Standardized image segmentation and feature extraction protocols, using the FreeSurfer processing software, developed by the ENIGMA consortium were used (http://enigma.ini.usc.edu/protocols/imaging-protocols/) to extract 153 features from regions of interest, including the volumes of 14 subcortical gray matter regions (bilateral nucleus accumbens, amygdala, caudate, hippocampus, pallidum, putamen, and thalamus) and the 2 lateral ventricles, cortical thickness and surface area from 68 cortical regions, and total intracranial volume (ICV). Segmentations were statistically examined for outliers and the FreeSurfer feature was excluded if it was >2.698 standard deviations away from the global mean. However, if a sample was a statistical outlier, but visual inspection showed that it was properly segmented, it was kept in the dataset.

### FreeSurfer brain age prediction model

We used a publicly available brain age model (https://www.photon-ai.com/enigma_brainage/) that was trained to predict age from 77 ((left+right hemisphere features)/2 and ICV) FreeSurfer features (for more detail, see (Han et al., 2019). Briefly, the Ridge Regression coefficients learned from 952 male and 1,236 female control subjects (aged 18-75 years) from the ENIGMA MDD working group were applied to the features of the current samples (N=285). Of note, NESDA was not part of the development of this model. The model’s generalization performance was assessed by calculating several metrics: a) the correlation between predicted brain age and chronological age, b) the amount of chronological age variance explained by the model (R_2_), c) the mean absolute error (MAE) between predicted brain age and chronological age, and d) Root Mean Squared Error (RMSE).

### Diagnostic ascertainment

Participants in the current study included control subjects (no lifetime history of psychiatric disorders) and patients with a current depression and/or current anxiety disorder (i.e. generalized anxiety disorder, panic disorder, social anxiety disorder) within a 6-month recency. The Composite International Diagnostic Interview (CIDI version 2.1) was used as a diagnostic instrument to ascertainment (Wittchen, 1994).

### Clinical assessment

We examined several clinical variables as predictors, including a) depressive symptoms as measured by the summary score of the Inventory for Depressive Symptoms (IDS) at time of scanning (Rush et al., 1996), but also b) three separate validated clusters of depressive symptoms (mood/cognition, somatic, and immunometabolic symptoms) (Wardenaar et al., 2010), c) anxiety symptoms as measured by the summary score of the Beck Anxiety Inventory (BAI) at time of scanning (Fydrich et al., 1992), d) cumulative childhood trauma index (Hovens et al., 2010) (before the age of 16) as measured by a childhood trauma interview, and e) recent negative life events in the past year as measured with the Brugha questionnaire (Brugha & Cragg, 1990). Within the patients only, we also investigated associations with: a) duration of symptoms, b) age of onset of illness, and c) antidepressant medication use (selective serotonin reuptake inhibitors (ATC code N06AB) and other antidepressants (ATC codes N06AF, N06AG, N06AX). See **Supplement** for full details.

### Somatic health assessment

Body Mass Index (BMI) was assessed during an interview by dividing a person’s weight (in kilogram [kg]) by the square of their height (in meter [m]). The number of self-reported current somatic diseases (heart disease, epilepsy, diabetes, osteoarthritis, cancer, stroke, intestinal disorders, ulcers, and lung-, liver-, and thyroid disease) for which participants received medical treatment was counted.

### Lifestyle assessment

Smoking status was expressed by calculating the number of cigarettes smoked per day. Alcohol consumption was expressed as the mean number of drinks consumed per week, measured by the AUDIT (Bush et al., 1998). Physical activity was assessed using the International Physical Activity Questionnaire (IPAQ) and expressed in total metabolic equivalent (MET) minutes per week (Craig et al., 2003).

### Biological stress assessment

We included predictors from three major biological stress systems: a) the immune-inflammatory system (C-reactive protein [CRP], Interleukin-6 (IL6), and tumor necrosis factor-α (TNF-α), b) the hypothalamic pituitary adrenal (HPA)-axis (cortisol awakening response [CAR] and evening cortisol), and c) the autonomic nervous system (ANS: heart rate, respiratory sinus arrhythmia [RSA] and pre-ejection period [PEP]). Details can be found in **Supplement**.

### Statistical analysis

All statistical analyses were performed using R version 3.5.3 (R Core Team, 2019). First, we used linear regressions to examine brain-PAD differences between the control and patient groups and tested brain-PAD associations with several clinical characteristics within the patients only (i.e. duration of symptoms, age of onset of illness, AD use). Second, we used separate linear regression models with brain-PAD as measured outcome and variables of interest as a predictor to explore and select significant contributors in all participants irrespective of diagnostic group. Finally, stepwise regression with forward selection was used to successively add significant contributors to an intercept-only model, starting with the variable that explained most variance and stopping if the model fit did not improve anymore. The best subset of variables leading to the best model fit (i.e. lowest Aikaike’s Information Criterion [AIC]) were selected to examine unique contributions to brain-PAD. Inflammatory predictors were log_e_-transformed due to highly skewed distributions and subsequently corrected for fasting status and anti-inflammatory medication use. ANS predictors were corrected for fasting status, heart medication use, and mean arterial blood pressure. HPA predictors were corrected for fasting status, awakening time, variable indicating whether it was a working day or not, and season. All biological stress markers >3*sd away from the mean were winsorized. Age, sex, education level (years), and two dummy variables for scan location were included as predictor variables in all models. Analyses were tested two-sided and findings were considered statistically significant at *p*<0.05. All b regression coefficients from all models may be interpreted as added brain aging in years in response to each unit increase of the predictor.

## Data Availability

Data sources are not available due to privacy issues, but we highly value scientific collaboration. Therefore, in principle, NESDA data are available to all scientific researchers working at non-commercial research organizations worldwide. Researchers can request either existing data for data analyses or bioanalysis. Please visit the online data overview for an extensive overview of the available data and NESDAs current output (www.nesda.nl). The used brain age prediction model can be found on https://www.photon-ai.com/enigma_brainage/.

https://www.photon-ai.com/enigma_brainage/

## ACKNOWLEDGEMENTS AND DISCLOSURES

The infrastructure for the Netherlands Study of Depression and Anxiety (https://www.nesda.nl) is funded through the Geestkracht program of the Netherlands Organization for Health Research and Development (Zon-MW, Grant number 10-000-1002) and is supported by participating universities and mental health care organizations (VU University Medical Center, GGZ inGeest, Arkin, Leiden University Medical Center, GGZ Rivierduinen, University Medical Center Groningen, Lentis, GGZ Friesland, GGZ Drenthe, Scientific Institute for Quality of Healthcare [IQ healthcare], and the Netherlands Institute of Mental Health and Addiction [Trimbos Institute]). LKMH and BWJHP were partly supported through the European Lifebrain consortium. BWJHP has received research funding (not related to the current paper) from Boehringer Ingelheim and Jansen Research. The other authors report no biomedical financial interests or potential conflicts of interest.

## Data availability

Data sources are not available due to privacy issues, but we highly value scientific collaboration. Therefore, in principle, NESDA data are available to all scientific researchers working at non-commercial research organizations worldwide. Researchers can request either existing data for data analyses or bioanalysis. Please visit the online data overview for an extensive overview of the available data and NESDA’s current output (www.nesda.nl). The used brain age prediction model can be found on https://www.photonai.com/enigma_brainage/.

### ARTICLE INFORMATION

Address correspondence to Laura K.M. Han, MSc, Department of Psychiatry, Amsterdam University Medical Centers, Location VUmc, Oldenaller 1, 1081 HJ Amsterdam, The Netherlands;

## REFERENCES

Aas, M., Elvsåshagen, T., Westlye, L. T., Kaufmann, T., Athanasiu, L., Djurovic, S., Melle, I., van der Meer, D., Martin-Ruiz, C., Steen, N. E., Agartz, I., & Andreassen, O. A. (2019). Telomere length is associated with childhood trauma in patients with severe mental disorders. Translational Psychiatry, 9 (1), 97.

Anatomical Therapeutic Chemical (ATC) Classification Index: Alphabetically Sorted According to Nonproprietary Drug Name ; Only ATC 5th Levels are Included. (n.d.). WHO Collaborating Centre for Drug Statistics and Methodology.

Bas-Hoogendam, J. M., van Steenbergen, H., Nienke Pannekoek, J., Fouche, J.-P., Lochner, C., Hattingh, C. J., Cremers, H. R., Furmark, T., Månsson, K. N. T., Frick, A., Engman, J., Boraxbekk, C.-J., Carlbring, P., Andersson, G., Fredrikson, M., Straube, T., Peterburs, J., Klumpp, H., Phan, K. L., … van der Wee, N. J. A. (2017). Voxel-based morphometry multi-center mega-analysis of brain structure in social anxiety disorder. NeuroImage. Clinical, 16, 678–688.

Belsky, D. W., Moffitt, T. E., Cohen, A. A., Corcoran, D. L., Levine, M. E., Prinz, J. A., Schaefer, J., Sugden, K., Williams, B., Poulton, R., & Caspi, A. (2018). Eleven Telomere, Epigenetic Clock, and Biomarker-Composite Quantifications of Biological Aging: Do They Measure the Same Thing? American Journal of Epidemiology, 187 (6), 1220–1230.

Besteher, B., Gaser, C., & Nenadić, I. (2019). Machine-learning based brain age estimation in major depression showing no evidence of accelerated aging. In Psychiatry Research: Neuroimaging (Vol. 290, pp. 1–4). https://doi.org/10.1016/j.pscychresns.2019.06.001

Bijl, R. V., & Ravelli, A. (2000). Current and residual functional disability associated with psychopathology: findings from the Netherlands Mental Health Survey and Incidence Study (NEMESIS). Psychological Medicine, 30 (3), 657–668.

Boldrini, M., Hen, R., Underwood, M. D., Rosoklija, G. B., Dwork, A. J., Mann, J. J., & Arango, V. (2012). Hippocampal angiogenesis and progenitor cell proliferation are increased with antidepressant use in major depression. Biological Psychiatry, 72 (7), 562–571.

Brainstorm Consortium, Anttila, V., Bulik-Sullivan, B., Finucane, H. K., Walters, R. K., Bras, J., Duncan, L., Escott-Price, V., Falcone, G. J., Gormley, P., Malik, R., Patsopoulos, N. A., Ripke, S., Wei, Z., Yu, D., Lee, P. H., Turley, P., Grenier-Boley, B., Chouraki, V., … Murray, R. (2018). Analysis of shared heritability in common disorders of the brain. Science, 360 (6395). https://doi.org/10.1126/science.aap8757

Brugha, T. S., & Cragg, D. (1990). The List of Threatening Experiences: the reliability and validity of a brief life events questionnaire. Acta Psychiatrica Scandinavica, 82 (1), 77–81.

Bush, K., Kivlahan, D. R., McDonell, M. B., Fihn, S. D., & Bradley, K. A. (1998). The AUDIT Alcohol Consumption Questions (AUDIT-C): An Effective Brief Screening Test for Problem Drinking. Archives of Internal Medicine, 158 (16), 1789–1795.

Caspi, A., Houts, R. M., Ambler, A., Danese, A., Elliott, M. L., Hariri, A., Harrington, H., Hogan, S., Poulton, R., Ramrakha, S., Rasmussen, L. J. H., Reuben, A., Richmond-Rakerd, L., Sugden, K., Wertz, J., Williams, B. S., & Moffitt, T. E. (2020). Longitudinal Assessment of Mental Health Disorders and Comorbidities Across 4 Decades Among Participants in the Dunedin Birth Cohort Study. JAMA Network Open, 3 (4), e203221.

Castrén, E., & Kojima, M. (2017). Brain-derived neurotrophic factor in mood disorders and antidepressant treatments. Neurobiology of Disease, 97 (Pt B), 119–126.

Cole, J., Franke, K., & Cherbuin, N. (2019). Quantification of the biological age of the brain using neuroimaging. In Healthy ageing and longevity. Biomarkers of Human Aging.

Cole, J. H. (n.d.). Multi-modality neuroimaging brain-age in UK Biobank: relationship to biomedical, lifestyle and cognitive factors. https://doi.org/10.1101/812982

Cole, J. H., & Franke, K. (2017). Predicting Age Using Neuroimaging: Innovative Brain Ageing Biomarkers. Trends in Neurosciences, 40 (12), 681–690.

Cousins, D. A., Aribisala, B., Nicol Ferrier, I., & Blamire, A. M. (2013). Lithium, gray matter, and magnetic resonance imaging signal. Biological Psychiatry, 73 (7), 652–657.

Craig, C. L., Marshall, A. L., Sjöström, M., Bauman, A. E., Booth, M. L., Ainsworth, B. E., Pratt, M., Ekelund, U., Yngve, A., Sallis, J. F., & Oja, P. (2003). International physical activity questionnaire: 12-country reliability and validity. Medicine and Science in Sports and Exercise, 35 (8), 1381–1395.

Debonnel, G., Saint-André, E., Hébert, C., de Montigny, C., Lavoie, N., & Blier, P. (2007). Differential physiological effects of a low dose and high doses of venlafaxine in major depression. The International Journal of Neuropsychopharmacology / Official Scientific Journal of the Collegium Internationale Neuropsychopharmacologicum, 10 (1), 51–61.

Eckel, R. H., Grundy, S. M., & Zimmet, P. Z. (2005). The metabolic syndrome. The Lancet, 365 (9468), 1415–1428.

Esposito, K., Chiodini, P., Colao, A., Lenzi, A., & Giugliano, D. (2012). Metabolic syndrome and risk of cancer: a systematic review and meta-analysis. Diabetes Care, 35 (11), 2402–2411.

Felger, J. C., Haroon, E., Patel, T. A., Goldsmith, D. R., Wommack, E. C., Woolwine, B. J., Le, N.-A., Feinberg, R., Tansey, M. G., & Miller, A. H. (2018). What does plasma CRP tell us about peripheral and central inflammation in depression? In Molecular Psychiatry. https://doi.org/10.1038/s41380-0180096-3

Ferrucci, L., Gonzalez-Freire, M., Fabbri, E., Simonsick, E., Tanaka, T., Moore, Z., Salimi, S., Sierra, F., & de Cabo, R. (2019). Measuring biological aging in humans: A quest. Aging Cell, e13080.

Franke, K., Gaser, C., Manor, B., & Novak, V. (2013). Advanced BrainAGE in older adults with type 2 diabetes mellitus. Frontiers in Aging Neuroscience, 5 (DEC), 1–9.

Franke, K., Ristow, M., & Gaser, C. (2014). Gender-specific impact of personal health parameters on individual brain aging in cognitively unimpaired elderly subjects. Frontiers in Aging Neuroscience, 6 (MAY), 1–14.

Fydrich, T., Dowdall, D., & Chambless, D. L. (1992). Reliability and validity of the Beck Anxiety Inventory. Journal of Anxiety Disorders, 6 (1), 55–61.

Gaser, C., & Franke, K. (2019). 10 years of BrainAGE as an neuroimaging biomarker of brain aging: What insights did we gain? Frontiers in Neurology, 10, 789.

Hägg, S., Belsky, D. W., & Cohen, A. A. (2019). Developments in molecular epidemiology of aging. Emerging Topics in Life Sciences, 3 (4), 411–421.

Han, L. K. M., Aghajani, M., Clark, S. L., Chan, R. F., Hattab, M. W., Shabalin, A. A., Zhao, M., Kumar, G., Xie, L. Y., Jansen, R., Milaneschi, Y., Dean, B., Aberg, K. A., van den Oord, E. J. C. G., & Penninx, B. W. J. H. (2018). Epigenetic Aging in Major Depressive Disorder. The American Journal of Psychiatry, 175 (8), 774–782.

Han, L. K. M., Dinga, R., Hahn, T., Ching, C., Eyler, L., Aftanas, L., Aghajani, M., Aleman, A., Baune, B., Berger, K., Brak, I., Filho, G. B., Carballedo, A., Connolly, C., Couvy-Duchesne, B., Cullen, K., Dannlowski, U., Davey, C., Dima, D., … Schmaal, L. (2019). Brain Aging in Major Depressive Disorder: Results from the ENIGMA Major Depressive Disorder working group. In bioRxiv (p. 560623). https://doi.org/10.1101/560623

Hovens, J. G. F. M., Wiersma, J. E., Giltay, E. J., Van Oppen, P., Spinhoven, P., Penninx, B. W. J. H., & Zitman, F. G. (2010). Childhood life events and childhood trauma in adult patients with depressive, anxiety and comorbid disorders vs. controls. Acta Psychiatrica Scandinavica, 122 (1), 66–74.

Howren, M. B., Lamkin, D. M., & Suls, J. (2009). Associations of depression with C-reactive protein, IL-1, and IL-6: a meta-analysis. Psychosomatic Medicine, 71 (2), 171–186.

Jansen, R., Verhoeven, J. E., Han, L. K. M., Aberg, K. A., van den Oord, E., Milaneschi, Y., & Penninx, B. (2020). An integrative study of five biological clocks in somatic and mental health. eLife (under review).

Jonsson, B. A., Bjornsdottir, G., Thorgeirsson, T. E., Ellingsen, L. M., Walters, G. B., Gudbjartsson, D. F., Stefansson, H., Stefansson, K., & Ulfarsson, M. O. (2019). Brain age prediction using deep learning uncovers associated sequence variants. Nature Communications, 10 (1), 5409.

Jylhava, J., Pedersen, N. L., & Hagg, S. (2017). Biological Age Predictors. EBioMedicine, 21, 29–36.

Kaufmann, T., van der Meer, D., Doan, N. T., Schwarz, E., Lund, M. J., Agartz, I., Alnæs, D., Barch, D. M., Baur-Streubel, R., Bertolino, A., Bettella, F., Beyer, M. K., Bøen, E., Borgwardt, S., Brandt, C. L., Buitelaar, J., Celius, E. G., Cervenka, S., Conzelmann, A., … Westlye, L. T. (2019). Common brain disorders are associated with heritable patterns of apparent aging of the brain. Nature Neuroscience, 22 (10), 1617–1623.

Kim, S., Myers, L., Wyckoff, J., Cherry, K. E., & Jazwinski, S. M. (2017). The frailty index outperforms DNA methylation age and its derivatives as an indicator of biological age. GeroScience, 39 (1), 83–92.

Kolenic, M., Franke, K., Hlinka, J., Matejka, M., Capkova, J., Pausova, Z., Uher, R., Alda, M., Spaniel, F., & Hajek, T. (2018). Obesity, dyslipidemia and brain age in first-episode psychosis. Journal of Psychiatric Research, 99 (vember 2017), 151–158.

Koutsouleris, N., Davatzikos, C., Borgwardt, S., Gaser, C., Bottlender, R., Frodl, T., Falkai, P., Riecher-Rössler, A., Möller, H. J., Reiser, M., Pantelis, C., & Meisenzahl, E. (2014). Accelerated brain aging in schizophrenia and beyond: A neuroanatomical marker of psychiatric disorders. Schizophrenia Bulletin, 40 (5), 1140–1153.

Kraus, C., Castrén, E., Kasper, S., & Lanzenberger, R. (2017). Serotonin and neuroplasticity--links between molecular, functional and structural pathophysiology in depression. Neuroscience and Biobehavioral Reviews, 77, 317–326.

Krueger, R. F., & Markon, K. E. (2006). Reinterpreting comorbidity: a model-based approach to understanding and classifying psychopathology. Annual Review of Clinical Psychology, 2, 111–133.

Le, T. T., Kuplicki, R. T., McKinney, B. A., Yeh, H.-W., Thompson, W. K., Paulus, M. P., & Tulsa 1000 Investigators. (2018). A Nonlinear Simulation Framework Supports Adjusting for Age When Analyzing BrainAGE. Frontiers in Aging Neuroscience, 10, 317.

Le, T. T., Kuplicki, R., Yeh, H. W., Aupperle, R. L., Khalsa, S. S., Simmons, W. K., & Paulus, M. P. (2018). Effect of Ibuprofen on BrainAGE: A Randomized, Placebo-Controlled, Dose-Response Exploratory Study. Biological Psychiatry: Cognitive Neuroscience and Neuroimaging, 1–8.

Lever-van Milligen, B. A., Verhoeven, J. E., Schmaal, L., van Velzen, L. S., Révész, D., Black, C. N., Han, L. K. M., Horsfall, M., Batelaan, N. M., van Balkom, A. J. L. M., van Schaik, D. J. F., van Oppen, P., & Penninx, B. W. J. H. (2019). The impact of depression and anxiety treatment on biological aging and metabolic stress: study protocol of the MOod treatment with antidepressants or running (MOTAR) study. BMC Psychiatry, 19 (1), 425.

Liang, H., Zhang, F., & Niu, X. (2019). Investigating systematic bias in brain age estimation with application to post-traumatic stress disorders. Human Brain Mapping, 10, 1.

Licht, C. M. M., de Geus, E. J. C., Zitman, F. G., Hoogendijk, W. J. G., van Dyck, R., & Penninx, B. W. J. H. (2008). Association between major depressive disorder and heart rate variability in the Netherlands Study of Depression and Anxiety (NESDA). Archives of General Psychiatry, 65 (12), 1358–1367.

Logue, M. W., van Rooij, S. J. H., Dennis, E. L., Davis, S. L., Hayes, J. P., Stevens, J. S., Densmore, M., Haswell, C. C., Ipser, J., Koch, S. B. J., Korgaonkar, M., Lebois, L. A. M., Peverill, M., Baker, J. T., Boedhoe, P. S. W., Frijling, J. L., Gruber, S. A., Harpaz-Rotem, I., Jahanshad, N., … Morey, R. A. (2018). Smaller Hippocampal Volume in Posttraumatic Stress Disorder: A Multisite ENIGMA-PGC Study: Subcortical Volumetry Results From Posttraumatic Stress Disorder Consortia. Biological Psychiatry, 83 (3), 244–253.

Molendijk, M. L., Bus, B. A. A., Spinhoven, P., Penninx, B. W. J. H., Kenis, G., Prickaerts, J., Voshaar, R. C. O., & Elzinga, B. M. (2011). Serum levels of brain-derived neurotrophic factor in major depressive disorder: state–trait issues, clinical features and pharmacological treatment. Molecular Psychiatry, 16 (11), 1088–1095.

Mottillo, S., Filion, K. B., Genest, J., Joseph, L., Pilote, L., Poirier, P., Rinfret, S., Schiffrin, E. L., & Eisenberg, M. J. (2010). The metabolic syndrome and cardiovascular risk a systematic review and meta-analysis. Journal of the American College of Cardiology, 56 (14), 1113–1132.

Murabito, J. M., Zhao, Q., Larson, M. G., Rong, J., Lin, H., Benjamin, E. J., Levy, D., & Lunetta, K. L. (2018). Measures of Biologic Age in a Community Sample Predict Mortality and Age-Related Disease: The Framingham Offspring Study. The Journals of Gerontology. Series A, Biological Sciences and Medical Sciences, 73 (6), 757–762.

Ning, K., Zhao, L., Matloff, W., Sun, F., & Toga, A. W. (2020). Association of relative brain age with tobacco smoking, alcohol consumption, and genetic variants. Scientific Reports, 10 (1), 10.

Pan, A., Keum, N., Okereke, O. I., Sun, Q., Kivimaki, M., Rubin, R. R., & Hu, F. B. (2012). Bidirectional association between depression and metabolic syndrome: a systematic review and meta-analysis of epidemiological studies. Diabetes Care, 35 (5), 1171–1180.

Penninx, B. W. J. H., Milaneschi, Y., Lamers, F., & Vogelzangs, N. (2013). Understanding the somatic consequences of depression: biological mechanisms and the role of depression symptom profile. BMC Medicine, 11, 129.

Profenno, L. A., Porsteinsson, A. P., & Faraone, S. V. (2010). Meta-analysis of Alzheimer’s disease risk with obesity, diabetes, and related disorders. Biological Psychiatry, 67 (6), 505–512.

Révész, D., Verhoeven, J. E., Milaneschi, Y., & Penninx, B. W. J. H. (2016). Depressive and anxiety disorders and short leukocyte telomere length: mediating effects of metabolic stress and lifestyle factors. Psychological Medicine, 46 (11), 2337–2349.

Roy-Byrne, P. P., Davidson, K. W., Kessler, R. C., Asmundson, G. J. G., Goodwin, R. D., Kubzansky, L., Lydiard, R. B., Massie, M. J., Katon, W., Laden, S. K., & Stein, M. B. (2008). Anxiety disorders and comorbid medical illness. General Hospital Psychiatry, 30 (3), 208–225.

Ruscio, A. M., & Khazanov, G. K. (2017). Anxiety and depression. The Oxford Handbook of Mood Disorders, 313–324.

Rush, A. J., Gullion, C. M., Basco, M. R., Jarrett, R. B., & Trivedi, M. H. (1996). The Inventory of Depressive Symptomatology (IDS): psychometric properties. Psychological Medicine, 26 (3), 477–486.

Santabárbara, J., Lopez-Anton, R., de la Cámara, C., Lobo, E., Gracia-García, P., Villagrasa, B., Bueno-Notivol, J., Marcos, G., & Lobo, A. (2019). Clinically significant anxiety as a risk factor for dementia in the elderly community. Acta Psychiatrica Scandinavica, 139 (1), 6–14.

Schmaal, L., Veltman, D. J., van Erp, T. G. M., Sämann, P. G., Frodl, T., Jahanshad, N., Loehrer, E., Tiemeier, H., Hofman, A., Niessen, W. J., Vernooij, M. W., Ikram, M. a., Wittfeld, K., Grabe, H. J., Block, A., Hegenscheid, K., Völzke, H., Hoehn, D., Czisch, M., … Hibar, D. P. (2015). Subcortical brain alterations in major depressive disorder: findings from the ENIGMA Major Depressive Disorder working group. Molecular Psychiatry, October 2014, 1–7.

Smith, S. M., Vidaurre, D., Alfaro-Almagro, F., Nichols, T. E., & Miller, K. L. (2019). Estimation of Brain Age Delta from Brain Imaging. https://doi.org/10.1101/560151

Steffener, J., Habeck, C., O’Shea, D., Razlighi, Q., Bherer, L., & Stern, Y. (2016). Differences between chronological and brain age are related to education and self-reported physical activity. Neurobiology of Aging, 40 (February), 138–144.

Van Gestel, H., Franke, K., Petite, J., Slaney, C., Garnham, J., Helmick, C., Johnson, K., Uher, R., Alda, M., & Hajek, T. (2019). Brain age in bipolar disorders: Effects of lithium treatment. The Australian and New Zealand Journal of Psychiatry, 53 (12), 1179–1188.

Verhoeven, J. E., Révész, D., Epel, E. S., Lin, J., Wolkowitz, O. M., & Penninx, B. W. J. H. (2013). Major depressive disorder and accelerated cellular aging: results from a large psychiatric cohort study. Molecular Psychiatry, May, 1–7.

Verhoeven, J. E., Révész, D., Van Oppen, P., Epel, E. S., Wolkowitz, O. M., & Penninx, B. W. J. H. (2015). Anxiety disorders and accelerated cellular ageing. British Journal of Psychiatry, 206 (5), 371–378.

Vreeburg, S. A., Hoogendijk, W. J. G., van Pelt, J., Derijk, R. H., Verhagen, J. C. M., van Dyck, R., Smit, J. H., Zitman, F. G., & Penninx, B. W. J. H. (2009). Major depressive disorder and hypothalamic-pituitary-adrenal axis activity: results from a large cohort study. Archives of General Psychiatry, 66 (6), 617–626.

Walker, E. R., McGee, R. E., & Druss, B. G. (2015). Mortality in mental disorders and global disease burden implications: a systematic review and meta-analysis. JAMA Psychiatry, 72 (4), 334–341.

Wardenaar, K. J., van Veen, T., Giltay, E. J., de Beurs, E., Penninx, B. W. J. H., & Zitman, F. G. (2010). Development and validation of a 30-item short adaptation of the Mood and Anxiety Symptoms Questionnaire (MASQ). Psychiatry Research, 179 (1), 101–106.

Wittchen, H. U. (1994). Reliability and validity studies of the WHO-Composite International Diagnostic Interview (CIDI): A critical review. Journal of Psychiatric Research, 28 (1), 57–84.

Wolf, E. J., Maniates, H., Nugent, N., Maihofer, A. X., Armstrong, D., Ratanatharathorn, A., Ashley-Koch, A. E., Garrett, M., Kimbrel, N. A., Lori, A., Va Mid-Atlantic Mirecc Workgroup, Aiello, A. E., Baker, D. G., Beckham, J. C., Boks, M. P., Galea, S., Geuze, E., Hauser, M. A., Kessler, R. C., … Logue, M. W. (2018). Traumatic stress and accelerated DNA methylation age: A meta-analysis. Psychoneuroendocrinology, 92, 123–134.

